# Viral Reactivation After Stroke: A Novel Model for Post-Stroke Depression

**DOI:** 10.1101/2025.04.02.25325132

**Authors:** Melanie Walker, Joseph R. Zunt, Michael R. Levitt, David M. Koelle

## Abstract

**Background:** Post-stroke depression (PSD) is a common and debilitating complication of acute ischemic stroke (AIS), yet its biological basis remains unclear. AIS induces immune dysregulation and blood–brain barrier (BBB) disruption, which may allow reactivation of latent viruses such as JC virus (JCV), a neurotropic polyomavirus that uses the serotonin receptor 5-HT2A to enter neurons involved in mood regulation.

**Methods:** We evaluated peripheral expression of host genes required for JCV entry, replication, and trafficking in three AIS transcriptomic datasets and assessed endothelial compromise using a complementary panel of BBB-associated genes. We then examined the anatomical relevance of viral entry receptors using publicly available brain expression datasets.

**Results:** AIS samples showed upregulation of host chaperone and trafficking genes, with concurrent downregulation of 5-HT2A. BBB-related profiles revealed reduced expression of structural junction proteins and increased expression of endothelial activation markers. Brain mapping localized high 5-HT2A expression to regions implicated in mood regulation.

**Conclusions:** These findings support a biologically plausible model in which AIS transiently enables JCV reactivation and CNS entry, particularly in serotonin-rich brain regions that may contribute to PSD pathogenesis.

## INTRODUCTION

Post-stroke depression (PSD) affects up to 50% of survivors of acute ischemic stroke (AIS),^1,2^ and significantly impairs functional recovery, increases healthcare utilization, and elevates long-term mortality.^3^ Although PSD has traditionally been viewed as a psychological reaction to disability or trauma, growing evidence suggests it arises from distinct biological mechanisms.^4–6^ Despite its high prevalence and impact, no definitive etiology or biomarker has been identified, and current treatment strategies remain limited.

AIS triggers a cascade of systemic and neurovascular changes, including immune dysregulation, inflammatory activation, and transient BBB disruption.^7–9^ These physiological shifts not only modulate injury response but also create a permissive environment for the reactivation of latent viral infections. One such pathogen is JCV, a prevalent human polyomavirus that establishes lifelong latency in peripheral tissues. By adulthood, approximately 60–80% of individuals are seropositive for JCV.^10,11^ Although typically asymptomatic, JCV can reactivate under conditions of immune suppression or systemic stress.^12–16^ Emerging evidence suggests it also reactivates and disseminates within the CNS in broader clinical contexts.^17–22^ In its most severe form, JCV reactivation causes progressive multifocal leukoencephalopathy (PML), a rare devastating CNS disease typically seen in immunocompromised individuals.^17,23,24^

JCV exhibits a strong neurotropism for serotonergic pathways.^25,26^ The virus primarily uses the serotonin receptor 5-HT2A (*HTR2A*) for cell entry^25,26^ — a receptor enriched in mood-regulating brain regions such as the prefrontal cortex, limbic system, and anterior cingulate cortex.^27^ These regions are implicated in the pathophysiology of depression.^28^ Similarly, 5-HT2B and 5-HT2C receptors have also been shown to support JCV entry via clathrin-mediated endocytosis *in vitro*.^29,30^ While initial viral attachment involves interactions with sialylated glycan structures on the cell surface,^30,31^ 5-HT2 receptors remain essential for internalization. These entry mechanisms may render serotonergic brain regions especially susceptible if JCV reactivates in the setting of stroke-related immune shifts or blood–brain barrier compromise. We hypothesize that AIS induces a systemic transcriptional environment permissive to JCV reactivation and neuroinvasion, which preferentially localizes to brain regions implicated in mood regulation and PSD.

## MATERIALS AND METHODS

This retrospective study analyzed publicly available human transcriptomic datasets of whole blood or peripheral blood mononuclear cells (PBMC) samples from AIS patients. The primary objective was to assess whether AIS induces a peripheral transcriptional profile consistent with biological permissiveness to JCV reactivation, including changes in host entry and replication factors. Secondary objectives included evaluating BBB-associated gene expression for signatures of endothelial disruption and mapping JCV- relevant host genes to brain regions implicated in mood regulation.

### Data Sources and Processing

Gene expression datasets were obtained from the National Center for Biotechnology Information’s Gene Expression Omnibus (GEO). Three datasets were selected based on relevance to AIS, availability of whole blood or PBMC profiles, and adequate sample size for group comparisons. These included GSE58294 (Affymetrix U133 Plus 2.0)^32^, GSE16561 (Illumina HumanRef-8 v3.0)^33^, and GSE37587 (Illumina HumanHT-12).^34^ GEO series matrix files and corresponding platform annotation files (GPL570, GPL6883, GPL10558) were downloaded and parsed using open-source Python libraries.^35^ Metadata were extracted from GEO annotations or inferred from sample order and documentation.

### Gene Panel Selection and Expression Analysis

A curated panel of human genes involved in JCV entry, trafficking, replication, and uncoating was assembled based on established functional annotations. JCV-relevant host factors included *HTR2A, TFRC, HSPA5, DNAJB11, STXBP1, STX4, DPM1, DPM2, DNAJC3, TOP1, and POLR2A*. A complementary set of genes related to BBB structure and function was added, including *CLDN5, OCLN, TJP1, PECAM1, CDH5, SLC2A1, VCAM1, ICAM1, SELE, SELP, PLVAP, LRP1, and MFSD2A*. Gene panels related to JC virus permissiveness and BBB function were assembled through literature review and keyword-based searches in Gene Ontology (GO),^36^ UniProtKB,^37^ and NCBI Gene databases.^38^ Search terms included combinations of functional descriptors such as “JC virus,” “polyomavirus,” “viral entry,” “serotonin receptor,” “vesicle trafficking,” “replication machinery,” and “protein uncoating” for the viral panel, and “tight junction,” “endothelial barrier,” “cell adhesion,” “transcytosis,” and “BBB permeability” for the BBB-related genes. Genes were retained if annotated functions were consistent across at least two sources. Functional groupings in Table 1 reflect shared roles in viral permissiveness or BBB regulation. Full gene names per HUGO Gene Nomenclature Committee^39^ are listed in Supplementary Table S1.

**Table 1.** JCV– and BBB–associated gene panels with functional categories.

| **Genes** | **Category** | **Function / Interpretation** |
| --- | --- | --- |
| ***HTR2A*** | Viral entry and receptor binding | 5-HT2A receptor mediating JCV entry |
| ***STXBP1, STX4*** | Vesicular trafficking / egress | Transport and exocytosis of viral components |
| ***POLR2A, TOP1*** | Viral replication machinery | Host polymerase and topoisomerase required for viral replication |
| ***HSPA5 (GRP78), DNAJB11, DNAJC3*** | Protein folding and uncoating | Endoplasmic reticulum chaperones supporting polyomavirus protein folding/uncoating |
| ***DPM1, DPM2*** | Glycosylation / viral maturation | Glycoprotein synthesis relevant to viral assembly |
| ***TFRC*** | Iron-dependent uptake | Iron transporter involved in glial JCV uptake |
| ***CLDN5, OCLN, TJP1*** | Tight junction structure | ↓ = compromised barrier integrity |
| ***PECAM1, CDH5, SLC2A1 (GLUT1)*** | Endothelial identity markers | ↓ = vascular injury or loss of barrier phenotype |
| ***VCAM1, ICAM1, SELE, SELP*** | Endothelial activation | ↑ = leukocyte adhesion and immune cell trafficking |
| ***PLVAP, LRP1, MFSD2A*** | Transcytosis / permeability control | ↑ PLVAP = fenestration; ↓ MFSD2A = disrupted transport selectivity |
| **Key:** Gene symbols follow standardized nomenclature from the HUGO Gene Nomenclature Committee (HGNC).<sup>36</sup> Functional groupings were assigned based on annotations from NCBI Gene, UniProt, and GeneCards, reflecting shared biological roles in JC virus infection or BBB integrity. Arrows indicate functional interpretation: ↓ suggests compromised structural or regulatory integrity and ↑ indicates upregulation or activation. |  |  |

Gene symbols were mapped to platform-specific probe IDs using each dataset’s annotation files. Probes with missing or non-numeric values were excluded. When multiple probes mapped to a gene, values were averaged. Expression values were extracted and z-score normalized across samples (by probe) to highlight relative variation. Heatmaps were generated using the seaborn package. Each heatmap reflects gene-level expression standardized to the dataset-wide mean.

We did not perform formal differential expression testing due to differences in preprocessing and platform scale. Instead, the analysis emphasized reproducibility of directionally consistent patterns across cohorts. Although control samples were available, heatmaps were not grouped by diagnosis to avoid artificial segmentation. Expression differences were assessed descriptively and summarized in Supplementary Tables S2 and S3.

### Brain Region Expression Mapping

To assess the neuroanatomical relevance of JCV-permissive host genes, RNA expression data were obtained from the Genotype-Tissue Expression (GTEx) Portal^40^ (v8.0), which provides bulk RNA-seq from postmortem human tissues. Transcript-per-million (TPM) values for *HTR2A, HSPA5, TFRC,* and *POLR2A* were extracted from seven GTEx brain categories: frontal cortex (BA9), anterior cingulate cortex (BA24), general cortex, hippocampus, amygdala, hypothalamus, and cerebellum. Protein-level regional expression and cell-type localization were cross-referenced using the Human Protein Atlas^41^ (v23.0), a publicly available resource which integrates immunohistochemistry and transcriptomic data. These data were used to evaluate the spatial distribution of viral entry and replication factors in mood-regulating brain regions, with relevance to PSD.^42,43^

### Statistical Analysis

Analysis focused on reproducibility and biological plausibility across independent cohorts. Given platform and sample variability, formal hypothesis testing (e.g., p-values, FDR correction) was not performed. Instead, z-score normalization and consistent directional changes were used to assess reproducible patterns.

## RESULTS

### JCV–Associated Host Gene Expression Following AIS

A total of 132 AIS samples and 47 controls were included across three datasets. In GSE58294, which includes whole blood collected at 3, 5, and 24 hours after AIS, 69 AIS and 23 control samples were available. Several genes showed altered expression in AIS samples compared to controls. Notably, probes for *STXBP1* and *TOP1* showed modest upregulation, while *HSPA5* and *HTR2A* were modestly downregulated. In GSE16561, a PBMC dataset with 39 AIS patients and 24 controls, a similar pattern was observed, with reduced *HTR2A* and increased *STXBP1, POLR2A,* and *DPM1*.

GSE37587, which includes blood and immune cell subtypes, contained 24 AIS samples and no controls, and showed further support for the hypothesis, with consistent upregulation of *TFRC, HSPA5*, and *DNAJB11*. These shifts were modest but reproducible across datasets and platforms. Of the 11 JCV-permissive genes, all were retained in the heatmaps except *STX4*, which lacked valid mapped probes in GSE16561. Expression patterns are shown in Figure 1A–C; gene-level summaries are in Supplementary Table S2.

**Figure 1.**
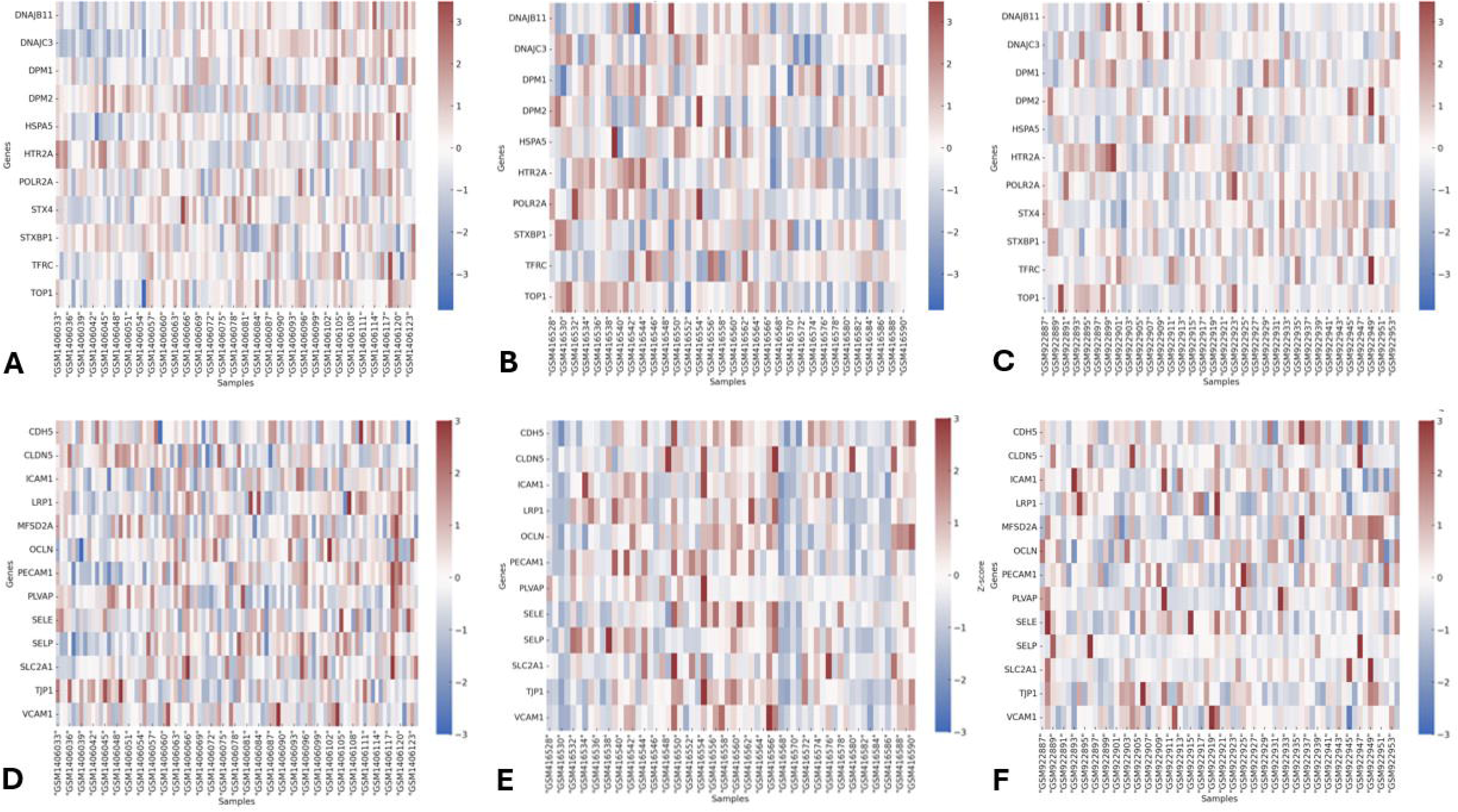
Expression of JC virus–associated and BBB–associated genes across three AIS datasets. Z-score normalized heatmaps displaying expression of JCV-permissive genes (Panels A–C) and BBB–associated genes (Panels D–F) across three publicly available gene expression datasets: GSE58294, GSE16561, and GSE37587. Gene panels are listed in Table 1. Heatmaps were generated from raw series matrix files using probe-to-gene mapping from corresponding annotation platforms. Each row represents a gene, and each column a sample. Color scale reflects Z-scores (range: –3 to +3), with red indicating above-average and blue indicating below-average expression relative to the dataset-wide mean. These expression patterns highlight peripheral transcriptional shifts consistent with viral permissiveness and concurrent BBB disruption in AIS.

### Blood-Brain Barrier Gene Expression After AIS

Across all three datasets, tight junction and endothelial integrity genes were consistently downregulated, while endothelial activation markers were upregulated in AIS samples relative to controls. GSE58294 and GSE16561 showed reduced expression of *CLDN5*, *PECAM1*, and *TJP1*, and increased *VCAM1, SELE*, and *PLVAP*. GSE37587 showed similar trends, particularly in *ICAM1, SELP*, and *PLVAP*. Of the 13 BBB-associated genes, all were retained in the heatmaps except *MFSD2A*, which lacked a mapped probe in GSE16561 but was recovered in GSE58294 and GSE37587 using platform-specific mappings. Heatmaps are shown in Figure 1D–F; directional summaries are in Supplementary Table S3.

### Brain Region Expression of JCV Host Genes

*HTR2A* was highly expressed in the frontal (31.6 TPM), anterior cingulate (30.3 TPM), and general cortices (29.7 TPM). *HSPA5* showed widespread expression with peak levels in the cerebellum (38.9 TPM), while *TFRC* and *POLR2A* were moderately to highly expressed in all brain regions. Data are summarized in Table 2; a barplot of *HTR2A* expression is in Supplementary Figure S1.

**Table 2.** Regional brain expression of JC virus host genes based on GTEx transcript-level data.

| Brain Region | *HTR2A* | *HSPA5* | *TFRC* | *POLR2A* |
| --- | --- | --- | --- | --- |
| Frontal Cortex (BA9) | 31.6 | 23.4 | 14.5 | 95.6 |
| Anterior Cingulate Cortex | 30.3 | 21.1 | 13.2 | 93.2 |
| Cortex (general) | 29.7 | 25.0 | 13.6 | 96.4 |
| Hippocampus | 24.9 | 24.2 | 14.8 | 94.7 |
| Amygdala | 25.3 | 23.1 | 13.5 | 93.1 |
| Hypothalamus | 22.0 | 21.7 | 15.7 | 92.3 |
| Cerebellum | 12.1 | 38.9 | 17.0 | 91.6 |
| Values represent median transcripts per million (TPM) for each gene across seven brain regions. Key: *HTR2A*: 5-HT2A serotonin receptor; *HSPA5*: heat shock protein family A (GRP78); *TFRC*: transferrin receptor; *POLR2A*: RNA polymerase II subunit A; GTEx: Genotype-Tissue Expression project. |  |  |  |  |

## DISCUSSION

This study proposes a model in which AIS induces a host environment permissive to JCV reactivation and potential neuroinvasion, with proclivity towards brain regions implicated in mood and PSD. Using three blood-derived transcriptomic datasets^44–46^, we detected a reproducible pattern of modest but directionally consistent changes in host genes required for JCV entry, replication, trafficking, and uncoating. AIS samples also showed expression signatures consistent with systemic endothelial activation and BBB compromise. While PML is classically associated with profound immunosuppression^17,23,24^, the conditions enabling JCV reactivation outside that context remain poorly defined. Our findings suggest AIS may represent a distinct, under- recognized permissive state, biologically different from PML but sufficient to induce a host environment favorable to JCV reactivation and potential neuroinvasion, with resultant clinical manifestations such as PSD.

The JCV host gene panel included *HTR2A*, which encodes the 5-HT2A serotonin receptor used for viral entry, along with genes involved in endoplasmic reticulum processing (*HSPA5*, *DNAJB11*), vesicular transport (*STXBP1*), and transcriptional machinery (*POLR2A*, *TOP1*). Across all three datasets, we observed reproducible upregulation of *HSPA5, STXBP1, TFRC,* and *DNAJB11* in AIS samples, while *HTR2A* was modestly downregulated. Though effect sizes were small, the consistency of these changes supports the plausibility of a stroke-induced peripheral transcriptional state permissive to viral reactivation. Importantly, JCV’s reliance on serotonergic receptors such as HTR2A gives it a unique neurotropic profile, distinguishing it from other latent viruses like herpes simplex virus (HSV), cytomegalovirus (CMV), and varicella-zoster virus (VZV), which utilize broader or non-serotonergic entry mechanisms,^47–49^ and raising the possibility of selective vulnerability in mood-regulating circuits.

To evaluate the potential for CNS entry, a curated BBB-associated gene panel was analyzed in the same AIS datasets. Genes related to tight junction integrity (*CLDN5, TJP1, PECAM1*) were consistently downregulated, while markers of endothelial activation (*VCAM1, ICAM1, SELE, PLVAP*) were upregulated. Although endothelial cells are rare in peripheral blood, these markers may reflect circulating inflammatory mediators or transcripts shed by activated endothelium, as described in other systemic inflammatory conditions.^50,51^ These patterns suggest AIS may trigger a peripheral immune state consistent with BBB disruption and increased leukocyte trafficking, creating a transient opportunity for viral entry.

Supporting this hypothesis, brain region expression data from GTEx and the Human Protein Atlas confirmed that *HTR2A* was highly enriched in the frontal cortex and anterior cingulate, both of which are associated with mood regulation and known to exhibit altered activity in PSD.^52^ *HSPA5*, *TFRC*, and *POLR2A* were also broadly expressed across cortical and limbic regions, reinforcing the potential for reactivated JCV to enter and replicate in serotonin-rich brain structures without causing overt cytopathology.

Together, these findings suggest a convergent model: AIS leads to transient immune dysregulation, downregulation of BBB structural components, and peripheral transcriptional shifts that enable reactivation of latent JCV. Once reactivated, JCV may preferentially affect serotonin-enriched brain regions via receptor-mediated entry, contributing to neuropsychiatric outcomes including PSD. This proposed pathway is mechanistically distinct from PML and may reflect a broader spectrum of pathology that is currently underrecognized.

This analysis has limitations. All transcriptomic data were derived from peripheral blood or PBMCs and may not fully capture intracellular changes in the CNS or endothelium. The observational design does not permit causal inference, and no direct evidence of viral reactivation or CNS infection was available in these cohorts. The absence of longitudinal follow-up or depression phenotyping also precludes clinical correlation. An additional limitation is the use of bulk RNA-seq, which does not distinguish the cellular source of observed expression changes. Single-cell RNA-seq was not available in these datasets. One dataset also lacked a control group, limiting comparative resolution. Despite these limitations, the reproducibility of transcriptional signatures and anatomical coherence of brain region expression support our model.

Based on guidelines originally designed for patients with PML^53^, an observational study based on this model is now underway. Future studies may incorporate paired blood and mood assessments, as well as evaluate JCV reactivation in related acute cerebrovascular conditions such as aneurysmal subarachnoid hemorrhage, which is also associated with a high incidence of subsequent depression.^54,55,56^ Our findings propose a biologically plausible model in which AIS may transiently enable JCV reactivation via a non-immunosuppressed pathway, expanding clinical relevance of beyond classical PML.

## Data Availability

All data produced in the present work are contained in the manuscript.

## ACKNOWLEDGEMENTS

N/A

