## Supplemental Tables and Figure for "Viral Reactivation After Stroke: A Novel Model for Post-Stroke Depression"

**Supplementary Table S1. Full gene names for JCV– and BBB-associated panels.**

| JCV-Associated Gene | Full Name |
|---|---|
| DNAJB11 | DnaJ Heat Shock Protein Family (Hsp40) Member B11 |
| DNAJC3 | DnaJ Heat Shock Protein Family (Hsp40) Member C3 |
| DPM1 | Dolichyl-Phosphate Mannosyltransferase Subunit 1 |
| DPM2 | Dolichyl-Phosphate Mannosyltransferase Regulatory Subunit |
| HSPA5 | Heat Shock Protein Family A (Hsp70) Member 5 |
| HTR2A | 5-Hydroxytryptamine Receptor 2A |
| LRP1 | LDL Receptor Related Protein 1 |
| PLVAP | Plasmalemma Vesicle Associated Protein |
| POLR2A | RNA Polymerase II Subunit A |
| STX4 | Syntaxin 4 |
| STXBP1 | Syntaxin Binding Protein 1 |
| TFRC | Transferrin Receptor |
| TOP1 | DNA Topoisomerase I |
| **BBB-Associated Gene** | **Full Name** |
| CDH5 | Cadherin 5 |
| CLDN5 | Claudin 5 |
| ICAM1 | Intercellular Adhesion Molecule 1 |
| MFSD2A | Major Facilitator Superfamily Domain Containing 2A |
| OCLN | Occludin |
| PECAM1 | Platelet And Endothelial Cell Adhesion Molecule 1 |
| SELE | Selectin E |
| SELP | Selectin P |
| SLC2A1 | Solute Carrier Family 2 Member 1 (GLUT1) |
| TJP1 | Tight Junction Protein 1 (ZO-1) |
| VCAM1 | Vascular Cell Adhesion Molecule 1 |
| **Key:** Gene symbols and full names are listed according to HUGO Gene Nomenclature Committee (HGNC) standards. | |

**Supplementary Table S2. Directional summary of JC virus–permissive gene expression in AIS vs control across three datasets.**

| | **Dataset** | | |
|---|---|---|---|
| **Gene** | **GSE58294** | **GSE16561** | **GSE37587** |
| *HTR2A* | ↓ | ↓ | ↓ |
| *HSPA5* | ↓ | – | ↑ |
| *TFRC* | – | ↑ | ↑ |
| *STXBP1* | ↑ | ↑ | – |
| *POLR2A* | – | ↑ | – |
| *DNAJB11* | – | – | ↑ |
| *TOP1* | ↑ | – | – |
| **Key**: ↑ = Higher in AIS; ↓ = Lower in AIS; – = No consistent change or not detected | | | |

**Supplementary Table S3. Directional summary of BBB-associated gene expression in AIS vs control. Summarizes probe-level directionality across all three datasets for the 13-gene BBB panel.**

| | **Dataset** | | |
|---|---|---|---|
| **Gene** | **GSE58294** | **Gene** | **GSE58294** |
| *CLDN5* | ↓ | ↓ | ↓ |
| *OCLN* | ↓ | – | – |
| *TJP1* | ↓ | ↓ | – |
| *PECAM1* | ↓ | – | ↓ |
| *CDH5* | ↓ | ↓ | – |
| *SLC2A1* | ↓ | – | ↓ |
| *VCAM1* | ↑ | ↑ | ↑ |
| *ICAM1* | ↑ | ↑ | ↑ |
| *SELE* | ↑ | ↑ | ↑ |
| *SELP* | ↑ | – | ↑ |
| *PLVAP* | ↑ | ↑ | ↑ |
| *LRP1* | – | – | ↑ |
| *MFSD2A* | ↓ | ↓ | – |
| **Key**: ↑ = Higher in AIS; ↓ = Lower in AIS; – = No consistent change or not detected | | | |

**Supplementary Figure S1. HTR2A expression across human brain regions (GTEx).**

Barplot shows transcript-level expression of the 5-HT2A receptor gene (HTR2A) across seven brain regions derived from the GTEx v8 dataset: frontal cortex (Brodmann area 9), anterior cingulate cortex (BA24), general cortex, hippocampus, amygdala, hypothalamus, and cerebellum. Expression was highest in the frontal cortex (31.6 TPM), anterior cingulate cortex (30.3 TPM), and general cortex (29.7 TPM), and lowest in the cerebellum (12.1 TPM), reflecting a >2.6-fold difference. The average expression across all regions was 25.1 TPM. These data support the hypothesis that JC virus, if reactivated and able to cross the BBB, may preferentially affect mood-related brain regions enriched in 5-HT2A expression.

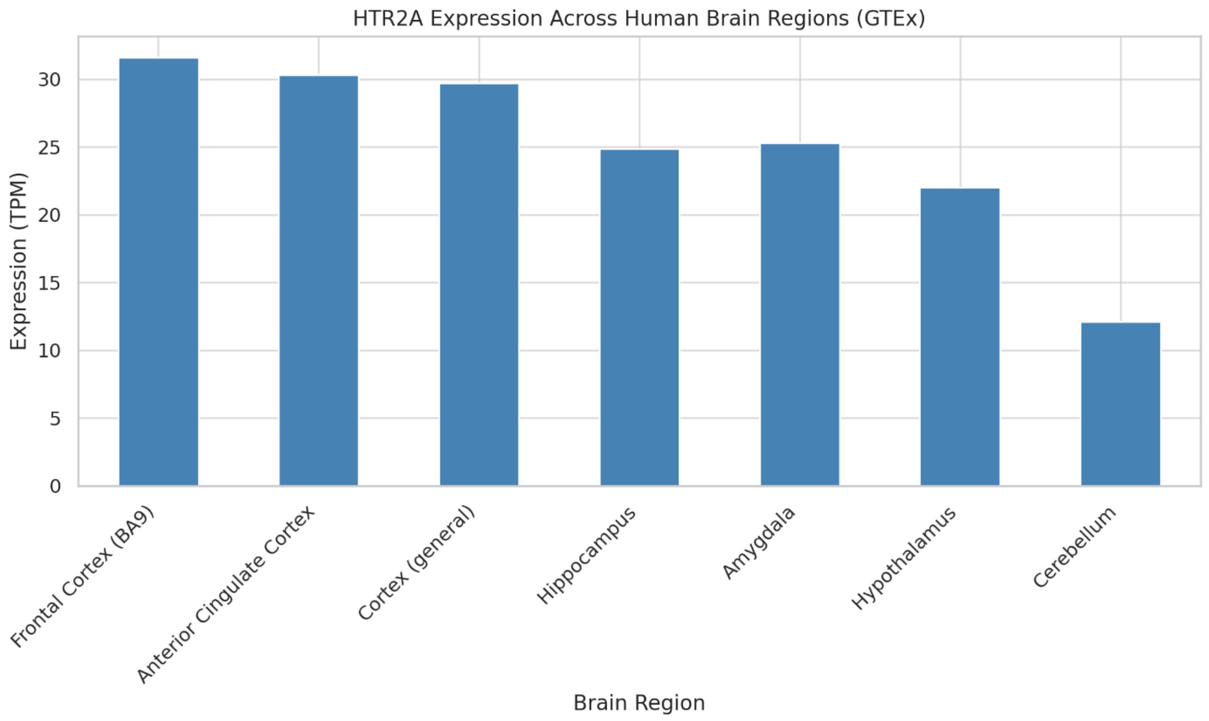
